# Identifying Medication-related Intents from a Bidirectional Text Messaging Platform for Hypertension Management: An Unsupervised Learning Approach

**DOI:** 10.1101/2021.12.23.21268061

**Authors:** Anahita Davoudi, Natalie S Lee, Thaibinh Luong, Timothy Delaney, Elizabeth L Asch, Krisda H Chaiyachati, Danielle L Mowery

## Abstract

**Background:** Free-text communication between patients and providers is playing an increasing role in chronic disease management, through platforms varying from traditional healthcare portals to more novel mobile messaging applications. These text data are rich resources for clinical and research purposes, but their sheer volume render them difficult to manage. Even automated approaches such as natural language processing require labor-intensive manual classification for developing training datasets, which is a rate-limiting step. Automated approaches to organizing free-text data are necessary to facilitate the use of free-text communication for clinical care and research.

**Objective:** We applied unsupervised learning approaches to 1) understand the types of topics discussed and 2) to learn medication-related intents from messages sent between patients and providers through a bidirectional text messaging system for managing participant blood pressure.

**Methods:** This study was a secondary analysis of de-identified messages from a remote mobile text-based employee hypertension management program at an academic institution. In experiment 1, we trained a Latent Dirichlet Allocation (LDA) model for each message type (inbound-patient and outbound-provider) and identified the distribution of major topics and significant topics (probability >0.20) across message types. In experiment 2, we annotated all medication-related messages with a single medication intent. Then, we trained a second LDA model (medLDA) to assess how well the unsupervised method could identify more fine-grained medication intents. We encoded each medication message with n-grams (n-1-3 words) using spaCy, clinical named entities using STANZA, and medication categories using MedEx, and then applied Chi-square feature selection to learn the most informative features associated with each medication intent.

**Results:** A total of 253 participants and 5 providers engaged in the program generating 12,131 total messages: 47% patient messages and 53% provider messages. Most patient messages correspond to blood pressure (BP) reporting, BP encouragement, and appointment scheduling. In contrast, most provider messages correspond to BP reporting, medication adherence, and confirmatory statements. In experiment 1, for both patient and provider messages, most messages contained 1 topic and few with more than 3 topics identified using LDA. However, manual review of some messages within topics revealed significant heterogeneity even within single-topic messages as identified by LDA. In experiment 2, among the 534 medication messages annotated with a single medication intent, most of the 282 patient medication messages referred to medication request (48%; n=134) and medication taking (28%; n=79); most of the 252 provider medication messages referred to medication question (69%; n=173). Although medLDA could identify a majority intent within each topic, the model could not distinguish medication intents with low prevalence within either patient or provider messages. Richer feature engineering identified informative lexical-semantic patterns associated with each medication intent class.

**Conclusion:** LDA can be an effective method for generating subgroups of messages with similar term usage and facilitate the review of topics to inform annotations. However, few training cases and shared vocabulary between intents precludes the use of LDA for fully automated deep medication intent classification.

## Introduction

Written communication is playing an increasing role in patient care. While secure platforms such as electronic health portals have historically provided the basis for such patient-provider communication, other streams of communication are rapidly developing. This is particularly true for management of chronic disease like hypertension, which has witnessed an explosion of mobile technologies, platforms, and programs supporting patient-provider communication outside the traditional office visit. These include wearables [1], social media platforms [2], and mobile apps [3].

These are all important developments. In the United States, hypertension, which is a leading cause of morbidity and mortality [4,5], affects approximately 45% of the adults, and is the most commonly diagnosed condition in the outpatient setting [6]. Smartphones are widely used by the general population and can play a valuable role in chronic disease management including chronic hypertension [7,8]. However, managing the sheer volume of clinical messages can be a source of clinician burnout due to technostress, time pressure, and workflow-related issues [9].

Automated approaches for classifying and processing patient-provider messages are promising for reducing clinician burden, particularly by harnessing the power of machine learning as in other industries [10]. Although several efforts have been made to classify intents from asynchronous communication platforms, few attempts have been made to develop methodologies that more efficiently filter and review messages to derive high-value intents; thereby, streamlining and automating the review of large corpora of text data [11–13]. We add to this body of literature by applying unsupervised learning methods to derive semantic topics from text-based digital health technologies e.g., text messaging systems, chatbots, and conversational agents. Our long-term goal is to develop robust methods for deriving patient-provider message intents from asynchronous, bi-directional communication platforms with minimal review effort. In this study, we apply unsupervised learning methods to 1) identify topics from messages generated between patients and providers and 2) assess the effectiveness of these methods to automatically identify subgroups of messages related to medication intents.

## Methods

In this University of Pennsylvania Institute Review Board-approved study (#834667), we retrospectively studied the text messages from adults enrolled in Penn Medicine’s Employee Hypertension Management Program (eHTN) from 2015 to 2019. To protect the privacy of our study participants, all data was deidentified using a text de-identification system called De-ID prior to study and analyses. In **Figure 1**, we outline our methodology and describe our analytical framework in subsequent sections.

**Figure 1.**
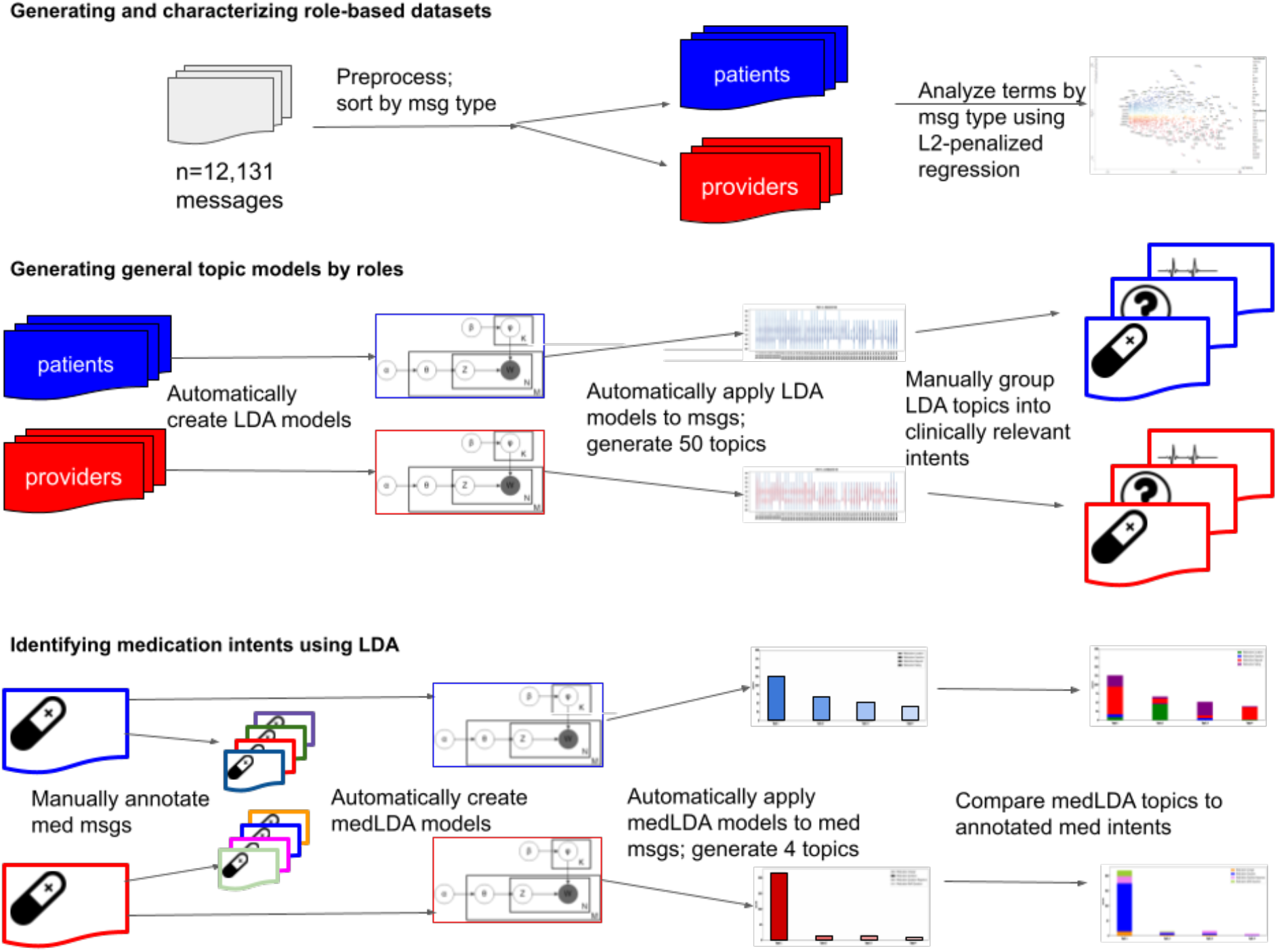
Study workflow and analysis. msg=messages; med=medications. LDA=Latent Dirichlet Allocation

### Employee Hypertension Management Program

The eHTN is a chronic disease management system dedicated to helping Penn Medicine employees with uncontrolled hypertension achieve and maintain controlled blood pressure (BP). Through the eHTN program, employees diagnosed with uncontrolled hypertension receive a prescription medication, a treatment plan for BP management, a BP cuff for home-based measurements, and access to a Penn Medicine registered nurse for free consultation. Our study period is from June 2015 through November 2019, when remote tracking of home BP readings, medication requests, and treatment counseling in the eHTN program was facilitated through a proprietary Health Insurance Portability and Accountability Act (HIPAA)-compliant, bi-directional text messaging application.

### Generating and characterizing role-based datasets

To automatically identify topical themes by roles of patient and providers, we classified messages into two datasets: inbound (patient-generated messages) and outbound (provider-generated messages). For each dataset, we removed messages that appeared to be automatic messages generated by the application, such as patient enrollment, e.g., “Registered to use the PROGRAM app” or calendar-events, e.g., “Calendar event created: Plan Check-In”. Among inbound messages, patients could report blood pressure readings as structured data elements (“Annotation: Pulse 76”) and add contextual information as free-text (e.g., “left arm”); for example, “Annotation: Pulse 95. Ran down the steps”. We removed all app-specific prefixes “Annotation:” from the message, but kept the remaining part of the text message in the model. Next, we preprocessed each message by reducing wordcase (“Meds” reduced to “meds”) and removing stop words (“of”, “the”). To better understand commonality and differences between the patient and provider messages, we analyzed and visualized individual words between datasets according to their frequency and informativeness. We computed and visualized the word frequencies according to their scaled L2-penalized regression coefficients using Scatterplot (https://spacy.io/universe/project/scattertext).

### Experiment 1, Part 1: Generating general topic models by roles

Next, we aimed to automatically learn topics dispersed within each message dataset. For each processed dataset (patient and provider independently), we applied Latent Dirichlet Allocation (LDA). LDA is an unsupervised, generative statistical model for learning subgroups of observations within a dataset based on similarities among observations [14]. We leveraged LDA to learn subgroups of messages with similar term usage and derive topics that might correlate to high-value intents hypothesized a priori (e.g., medication reorders, appointment scheduling requests, etc.). To learn useful LDA models, parameters for each model were set as follows: alpha was set as asymmetric to ensure that document-topic density results in more specific topic distributions per document; beta was set as symmetric to ensure topic-word density results in less specific word distributions per topic. This means that topics should be precise, but the words that compose a topic can be diverse and comprise of most words in the corpus. We limited the number of topics to 50 after observing that the exact composition of terms listed across models with topics over 50 were identical with near zero weights suggesting that LDA was unable to identify any additional distinct topics beyond 50 topics. In LDA models, each message has a probability associated with each derived topic. For each message, the probabilities of each topic sum to 1; therefore, a message could have one or more significant subtopics. We defined a *main topic* as the topic with the highest probability; *a significant subtopic* was defined as any topic with a probability equal or greater than 0.20. Note, that a main topic can be less than 0.20 because some messages may or may not have a significant topic, i.e., above 0.20 probability. For each message, we identified both main and significant subtopics. Among each type of message (patient and provider), we report the following distributions: messages with multiple topics, main topics across messages, and significant subtopics associated with each main topic.

### Experiment 1, Part 2: Validating LDA-derived topics

To explore the usability of LDA-derived topics, we manually annotated a subset of messages.The codebook for manual annotation was loosely based on an annotation schema from prior work on clinician-patient text communication [15], refined by review of over 1200 text messages exchanged in the program between October 1, 2020 and January 31, 2021 (TB, NL) conducted for a separate internal pilot study, and further refined based on iterative discussion and review of the study text data (TB, NL, DM) [15]. We attempted to apply this schema to each LDA-derived topic, but determined that the patient messages still contained heterogeneity (1 topic does not correlate to a single intent) and variability of intents. Given these factors,, we focused our annotation efforts solely on those messages which had been classified as having a single LDA intent, occurred frequently, and appeared clinically useful; we chose medication-related intents for this last category, given the clinical importance of identifying medication related communication. We narrowed our focus to these messages for Experiment 2.

### Experiment 2, Part 1: Identifying medication intents using LDA

For each subtopic, we ranked all messages assigned to the subtopic based on its probability for review and classification. Each of three reviewers (authors TL, DM, NL) manually reviewed the ranked messages as three distinct message sets expanding intent categories as described above. Within each subtopic, each author identified and classified each relevant message according to a single medication intent; any messages that were deemed to have more than one intent were excluded from review, with the exception of messages where the second intent was a pleasantry. All medication messages were reviewed among the team to resolve disagreements through consensus. For all messages annotated with a single medication intent, we generated another medication-specific LDA model (medLDA) and attempted to reclassify the resulting messages according to the set k topics (k=4 for both patient and provider messages, based on manual annotation). We reported the distribution of classified medication intents within each medLDA topic.

### Experiment 2, Part 2: Visualizing sublanguage of medication intents

We aimed to automatically capture the sublanguage of medication intents by identifying the most significant language features for each medication intent. To identify lexical features, each text message was preprocessed using spaCy by removing stop words, reducing case and encoding n-grams. We applied TF-IDF (term-frequency/inverse document frequency) to extract the most informative lexical features. To identify semantic features, we encoded the named entities of *problems, treatments*, and *tests* using the i2b2 Named Entity Recognition (NER) model from the STANZA python package [16]. To standardize medication-related details, we encoded RxNorm categories and semantic medication categories of *drug name, strength, route, frequency, form, dose amount, intake time, duration, dispense amount, refill, necessity* using the MedEx package [17]. Examples of semantic features can be found in **Table 1**. The stop words have been removed in the preprocessing step and question marks are shown by the “question” phrase in the WordClouds. Within each subclass, we computed the word (n=1-3 word) frequencies and identified the most informative words using chi-square feature selection. We selected the lexical and semantic features (n-grams, STANZA NERs, and MedEx) most significantly associated with each medication intent (p<0.05).We applied a log-10 transform to each of the p-values so features with higher significance would appear larger when we visualized the most significant features for each medication intent using WordClouds.

**Table 1.** Examples for lexical and semantic features.

| Package Type | Category | Example |
| --- | --- | --- |
| STANZA | Problem | “Musinex, proair inhaler and delsym for **cough**” |
|  | Test | “Took meds between 8:30 and 9:00 am after my **MRI**” |
|  | Treatment | “**Meds** taken late” |
| MedEx | DPN | “Sorry Lisinopril was stopped **HCTZ** 25 mg added / daily Metoprolol decreased to 50 mg / daily.” |
|  | Drug Ingredient (DIN) | “Sorry **Lisinopril** was stopped HCTZ 25 mg added / daily **Metoprolol** decreased to 50 mg / daily.” |
|  | Drug Brand Name (DBN) | “Hi have only 11 tablets of **amlodipine** left. Are you able to issue me a new prescription?” |
|  | Drug Dose Form (DDF) | “Dr doubled my Lisinopril and removed the water **pill** See OV” |
|  | Dose (DOSE) | “Sorry Lisinopril was stopped HCTZ **25 mg** added / daily Metoprolol decreased to **50 mg** / daily.” |
|  | Dose amount (DOSEAMT) | “Hi have only **11 tablets** of amlodipine left. Are you able to issue me a new prescription?” |
|  | Frequency (FREQ) | “Sorry Lisinopril was stopped HCTZ 25 mg added / **daily** Metoprolol decreased to 50 mg / **daily**.” |
|  | Route (RUT) | “Lossrtan is giving me SOB chest pressure on **inhalation** is this l? Happens about 20 min after I take it and lasts thru day.” |
|  | Duration (DRT) | “Lossrtan is giving me SOB chest pressure on inhalation is this l? Happens about **20 min** after I take it and lasts thru day.” |
|  | DOSE UNIT | “Good Morning can you call me in a refill for my bp **pills** please?” |

## Results

In this retrospective, observational study, we studied 253 participants who were enrolled in the eHTN program. Of the patients who participated, 96% (243/253) were actively engaged in the program sending at least 1 inbound message. In **Table 2**, of the total 12,131 messages collected, 47% (n=5,689) of messages were generated by patients; 53% (n=6,442) of messages were generated by providers.

**Table 2.** Statistics according to patient and provider message dataset.

| Message Type<br>(mean +/- stdev) [range] | Patient messages<br>(n=5,689 messages) | Provider messages<br>(n=6,442 messages) |
| --- | --- | --- |
| Words per message | 17.01+/- 23.40 [1, 1176] | 26.75+/- 28.79 [1, 299] |
| Sentences per message | 2.71+/- 2.02 [1, 68] | 3.11+/- 2.11 [1, 15] |
| Messages per user | 23.84 +/- 26.22 [1, 232] | 521.83 +/-1588.2 [1, 5558] |

In **Figure 2**, we show the relationship between terms and dataset. Within the patient messages (blue), we observed terms with higher coefficients and higher frequency including temporal expressions (“morning”, “evening”, “tonight”, “gm (good morning)”, “hr”), medical terms (“rx”, “pulse”), and confirmations (“okay”, “thx”). Within the provider messages (red), we observed terms with lower coefficients and higher frequency including salutations (“mrs”, “mr”), directive verbs (“check record”, “sign”, “send”, “confirm”, “recheck”, “look”) and positive and negative sentiment (“nice”, “great”, “worry”).

**Figure 2.**
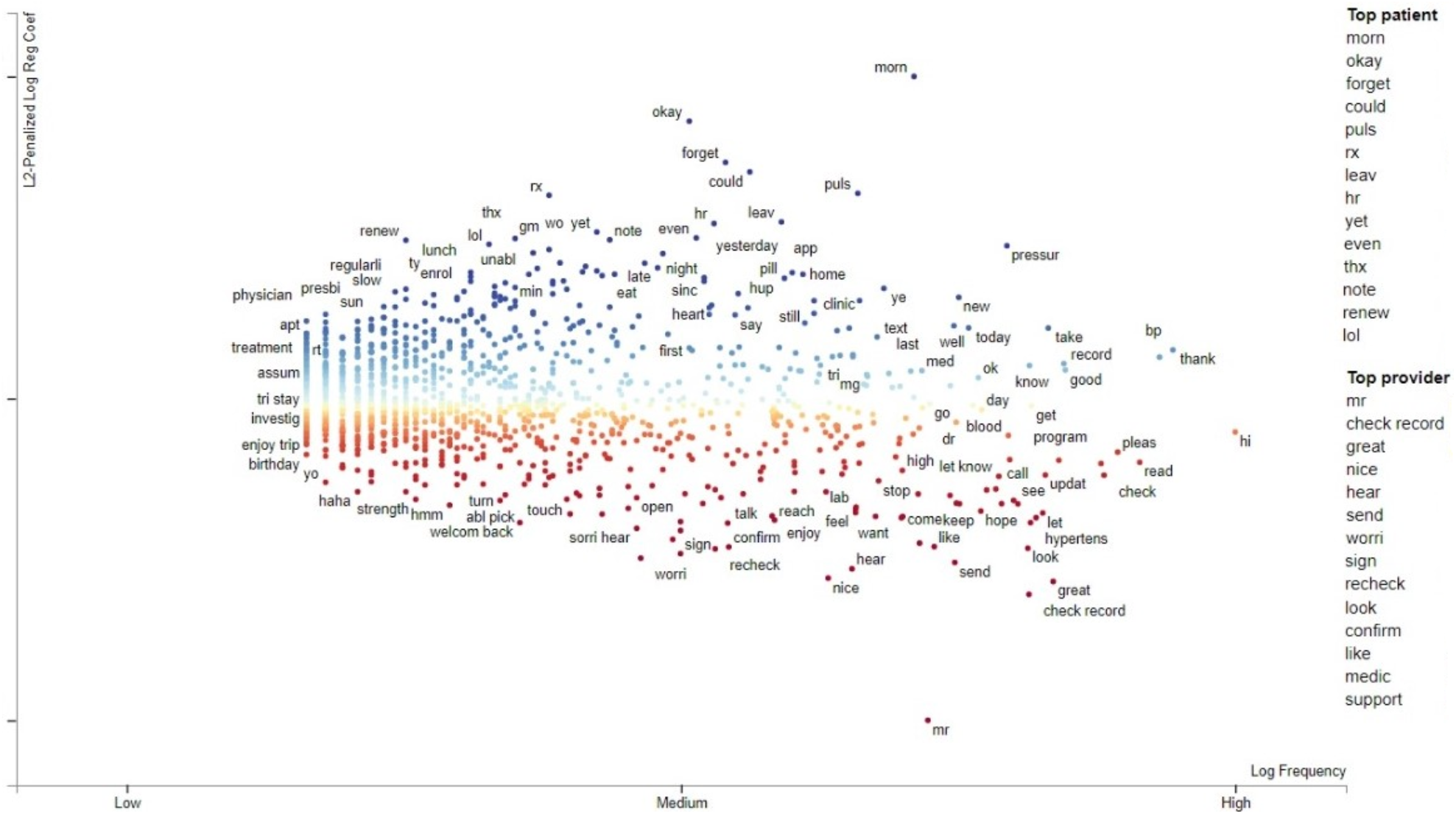
Characteristics of messages with scatterplot image of patient (blue) versus provider (red) word frequency.

In **Figure 3**, we show the distribution of topics for patient and provider messages based on (**A**) major topics, (**B**) shared significant topics, and (**C**) significant subtopics within a topic. The significant topic is defined where the probability of that topic within a message is > 0.2. Among the patient messages, the majority of messages defined by predictive “terms” occur within topics 1 (n=1117 messages; “thank, ok, great, nice”), 17 (n=412 messages; “pulse, mg, daili, take, tab, dose, losartan, amlopidin”), and 7 (n=395 messages; “bp, read, hi, record, check, today”); among the provider messages, the majority of messages occur within topics 47 (n=1249 messages; “record, please, bp, update, read, hi, check”), 12 (n=665 messages; “good, look, work, bp, keep, great”), and 42 (n=419 messages; “call, schedul, hi, appt, please, appoint, come, see follow”). We observed potential topics within each patient and provider groups. Some messages consist of 1 to 4 topics based on our model. Among the patient messages, most messages have 1 significant topic (n=2851 messages) followed by 2 (n=1893 messages) and 3 (n=564 messages) significant topics. Similarly, among the provider messages, most messages have 1 significant topic (n=3311 messages) followed by 2 (n=2466 messages) and 3 (n=503 messages) significant topics. Within the patient and provider groups, messages range from having 0 up to 4 significant topics. Among the patient messages within each topic, 5 topics (topics 17, 34, 43, 44, 46) appear to have 1 additional significant topic. Among the provider messages within each topic, 6 topics (topics 22, 29, 30, 31, 33, 38) appear to have 1 additional significant topic. The term list and associated probabilities within each aforementioned topic can be found in the **Supplement - LDA topics**.

**Figure 3.**
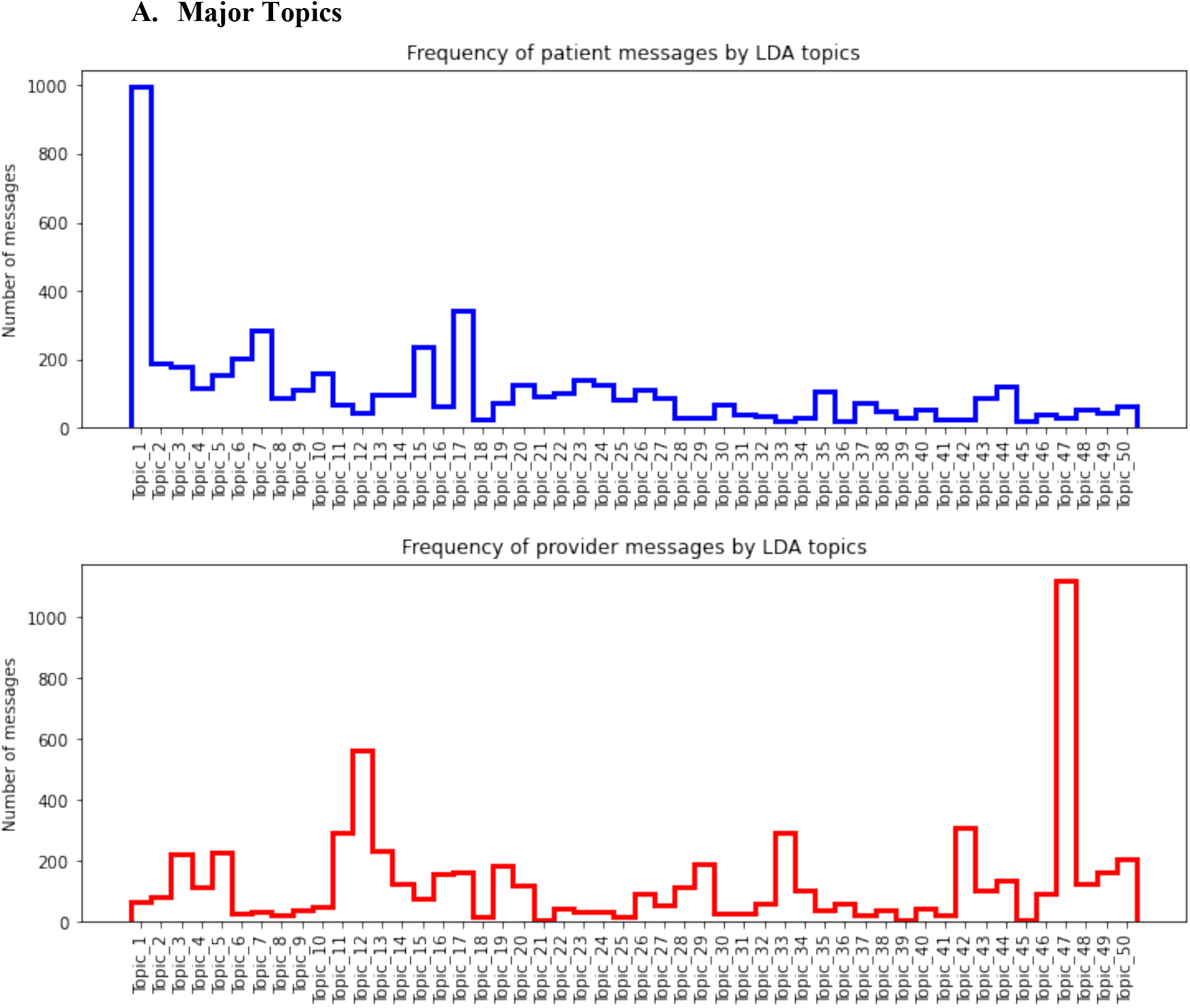
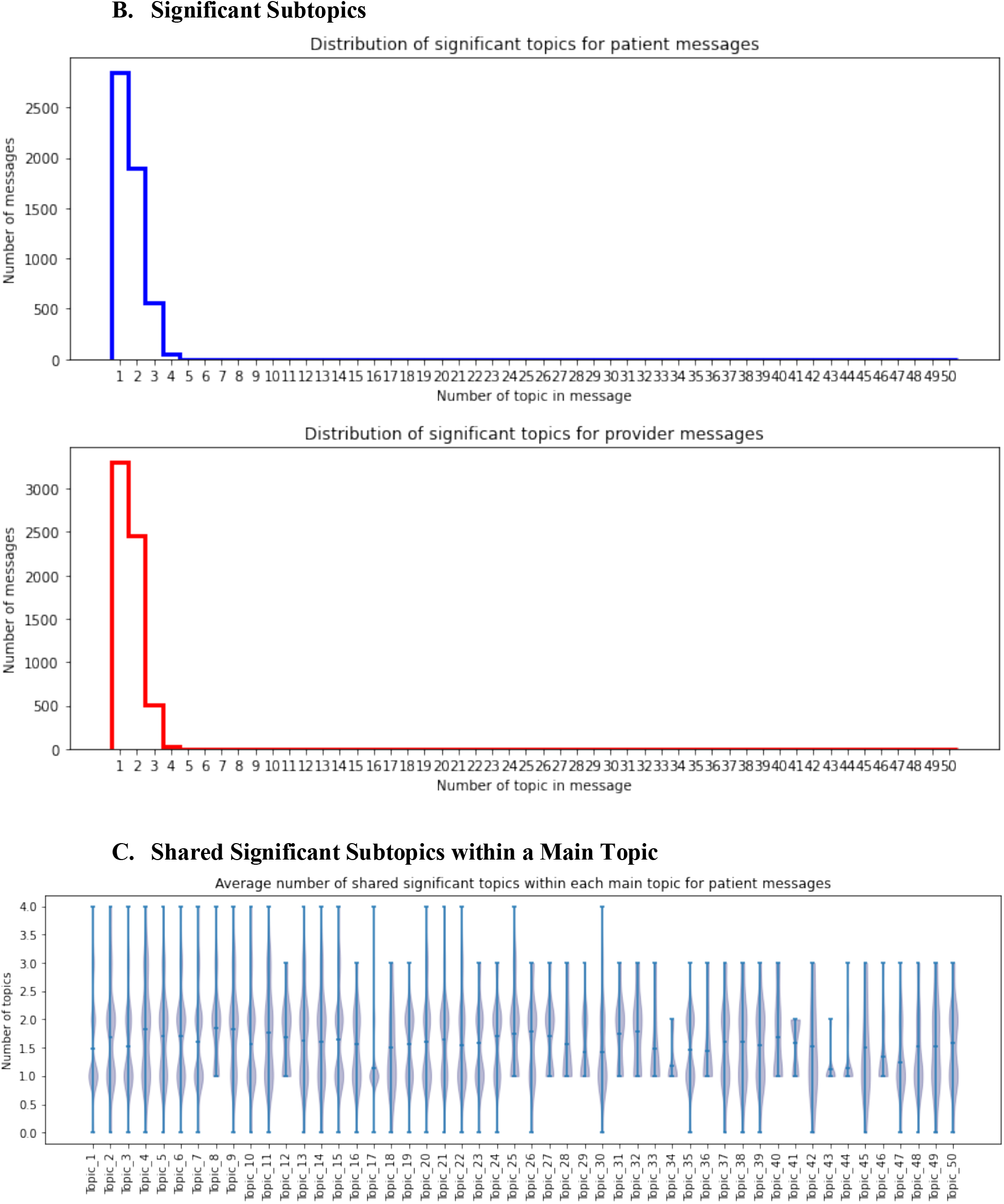
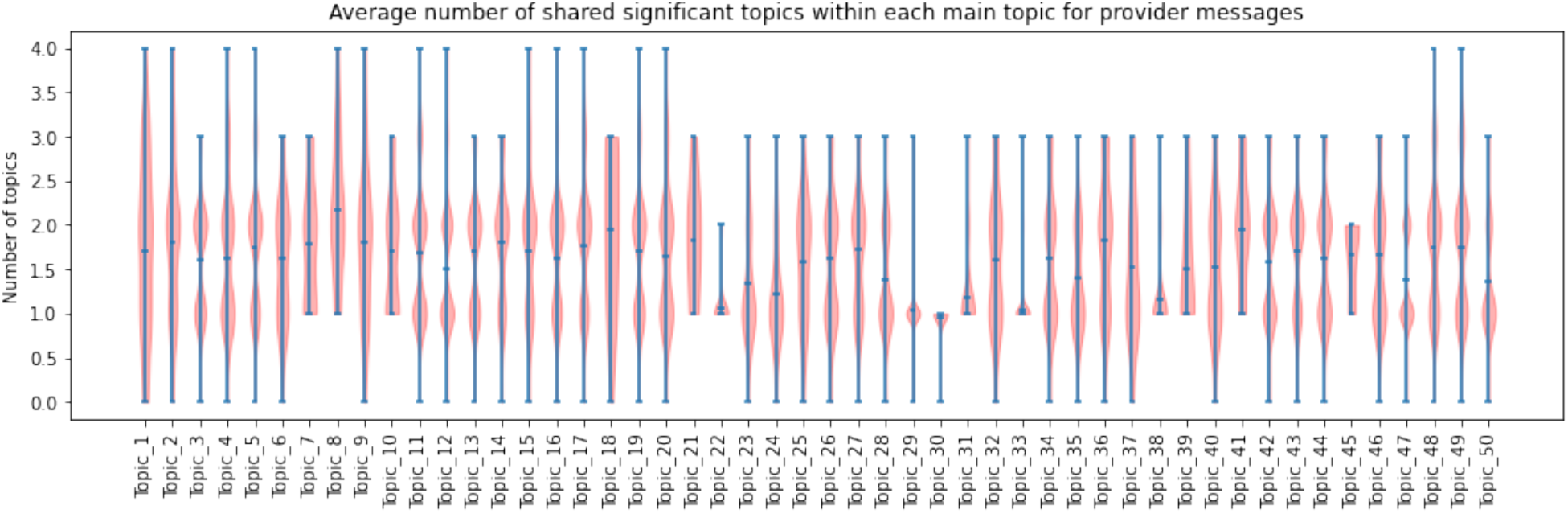
Distribution of messages according to major topics (**A**), shared significant subtopics (**B**), and significant subtopics within each main topic (**C**).

In **Table 3**, among the 282 patient medication messages, the majority of messages were *medication request* (47.5%; 134/282) followed by *medication taking* (28.0%; 79/282) and *medication location* (19%; 54/282); among the 252 provider medication messages, the majority of messages were *medication question* (68.7%; 173/252) followed by *medication question response* (16.3%; 41/252).

**Table 3.**
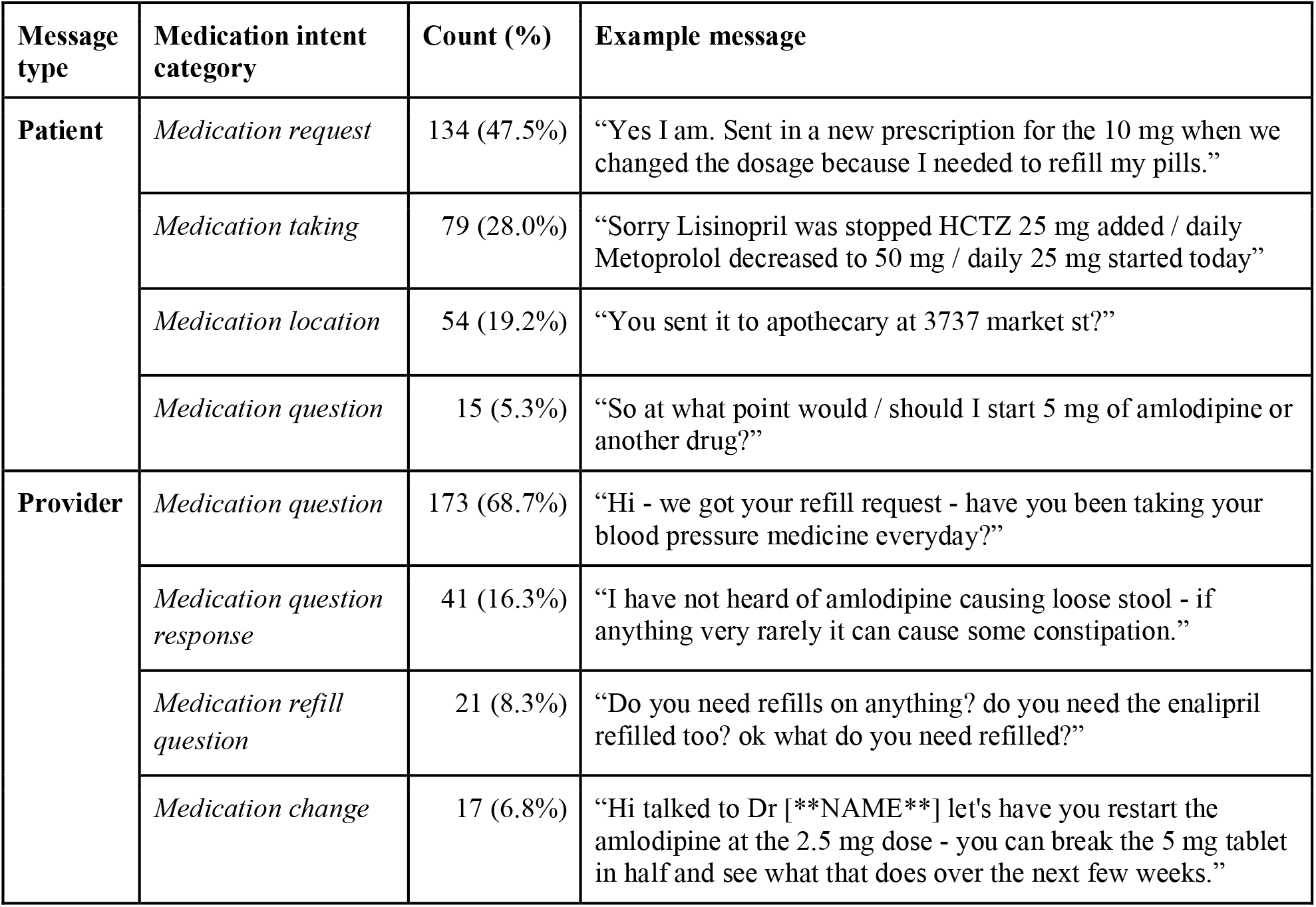
Distribution of medication intent categories with examples from patient and provider messages

In **Figure 4**, we depict the outcomes of the medLDA experiment for both patient and provider messages. Among the patient messages manually classified according to medication intent and automatically classified within a topic, we observe a majority medication intent within each topic: *medication request* (topic 1), *medication location* (topic 2), *medication taking* (topic 3), and *medication request* (topic 4). Among the provider messages, the majority of medication intents within each topic were *medication question* (topics 1 and 2) and *medication question response* (topics 3 and 4). The term list and associated probabilities within each topic can be found in the **Supplement - medLDA topics**.

**Figure 4.**
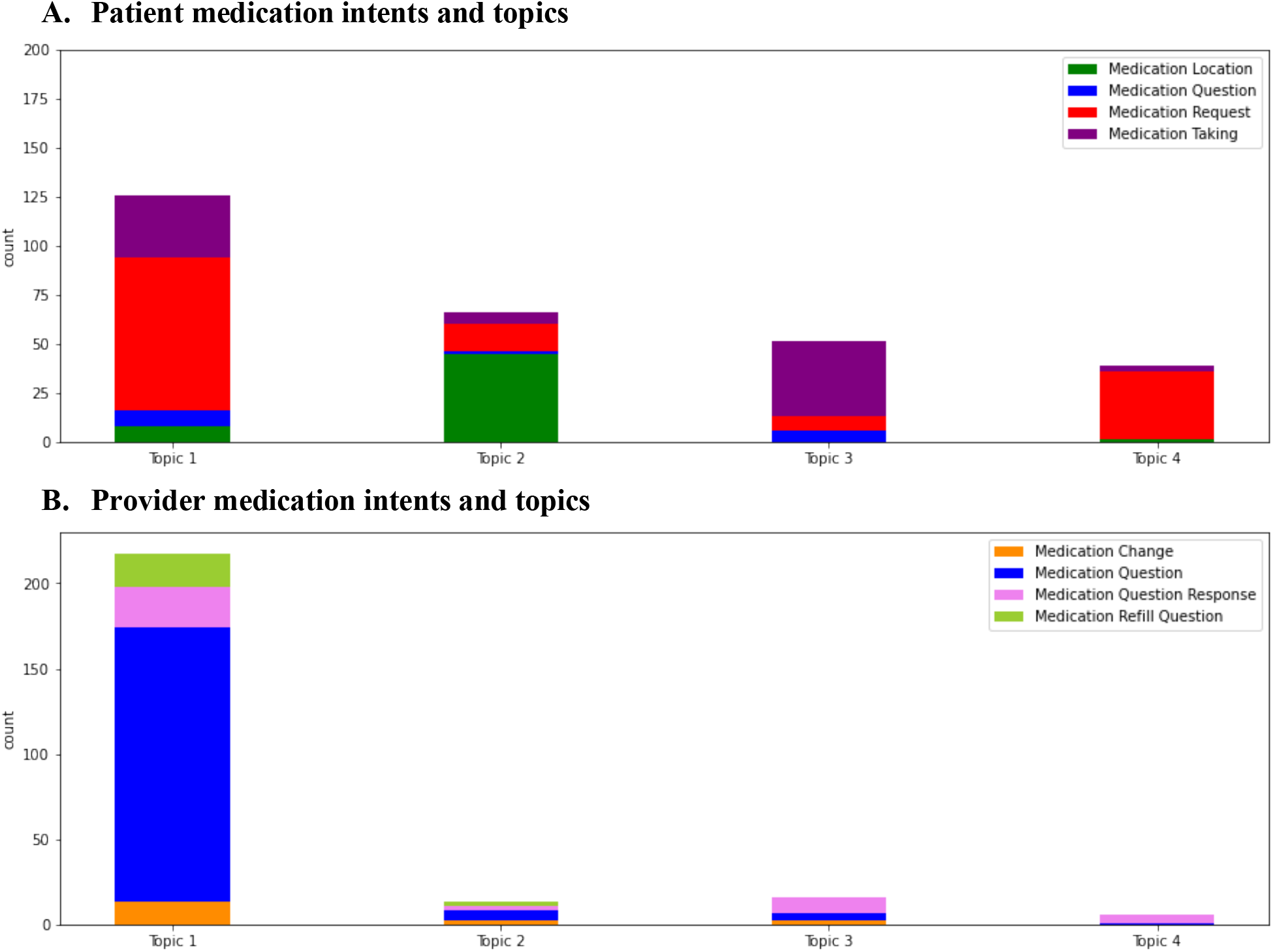
Distribution of medication intents among patient (A) and provider (B) messages after medLDA.

In **Figure 5**, we observe lexical and semantic features associated within each medication intent category according to patient (**A**) and provider (**B**) dataset. Among the patient medication intents, for *medication locations*, terms associated with drug dispensaries (“pharmacy”, “apothecary”), hospitals (“hup”= Hospital of the University of Pennsylvania), and street locations (“Spruce”, “Market”) are common. For *medication question,* we observed semantic types associated with MedEx drug names (DPN, DBN: Drug Brand Name, DDF: Drug Dose Form, DIN: Drug Ingredient), course (“start”, “stop”), use of a question mark and STANZA problem entity. *Medication request* and *medication taking* commonly include MedEx categories (DOSE and FREQ), terms for refills and verbs (“need”, “taking”) and STANZA treatment entities. Among the provider medication intents, for *medication change*, terms associated with temporal expressions (“tomorrow”, “week”), MedEx categories (DBN, DOSE), and program references are common. Among *medication question* and *medication refill question,* we observed terms associated with refill requests (“refill”, “refilled” and “need”), use of a question mark, and STANZA treatment entity. Among *medication question response*, we observe terms associated with references and change (“baseline” and “increasing”) as well as side effects.

**Figure 5.**
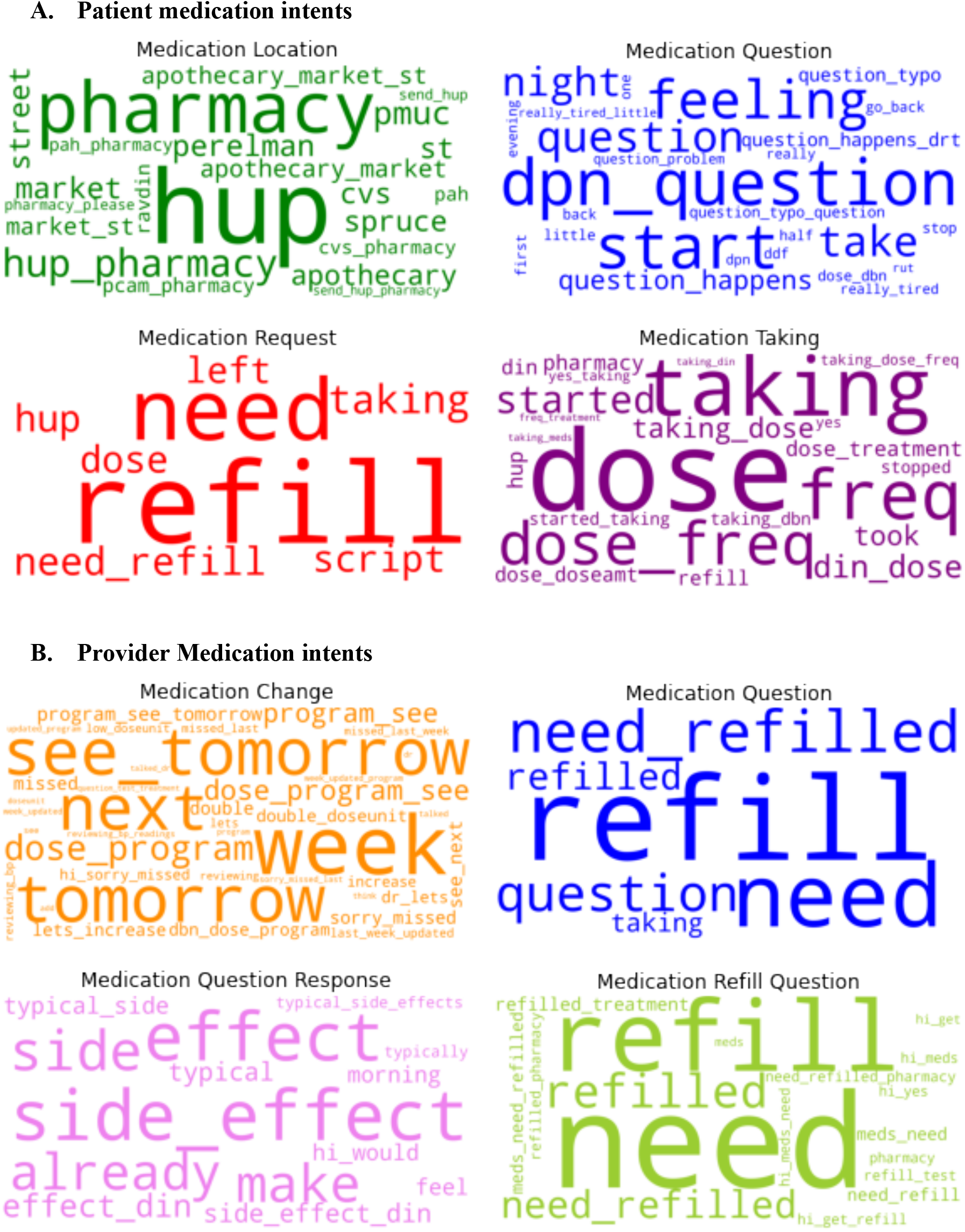
Word clouds of most informative terms associated with each medication intent class for patient and provider messages.

## Discussion

We developed and applied an unsupervised method, LDA, to facilitate the review and derivation of patient and provider intents from a large dataset of messages produced using an asynchronous, bi-directional communication platform. We learned LDA can be leveraged to identify subgroups of messages for systematic review generating insights into patient and provider information needs, albeit with some limitations.

To detect distinct word usage between patients (inbound) and providers (outbound), we applied a data-driven approach using single word frequencies scaled by L2-penalized regression coefficients and visualized word differences between messages categorized by role. We learned that patients often use temporal expressions (“morning”, “evening”) to initiate requests as well as medical terms (“rx”, “pulse”), to communicate medication and BP reporting, and confirmations (“okay”, “thx”) to convey information understanding, respectively. We also learned that providers express salutations (“mrs”, “mr”) to initiate communication with patients, directive verbs (“check record”, “sign”, “send”, “confirm”, “recheck”, “look”) to instruct patients to take actions important for informing clinical decisions by the care team, and positive and negative sentiment (“nice”, “great”, “worry”) to encourage patients to continue program engagement.

These initial insights informed the decision to develop LDA models based on roles -- patient and provider -- to identify potentially distinct topics among messages. When developing LDA models by patient and provider messages, the majority of messages occurred within a few prominent major topics. Among the most common topics from patient-generated messages, we observed topics with terms indicative of BP checking/reporting (topic 47; “bp, check”), blood pressure reporting encouragement (topic 12; “good, work, keep, great”), and appointment scheduling (topic 42; “call, appt, come”). Among the most common topics from provider-generated messages, we observed topics with terms suggestive of confirmation/gratitude (topic 1; “thank, nice”), medication adherence (topic 17; “mg, tab, dose”), and BP reporting (topic 7; “bp, record, check”). In a few topics, we observed a single intent which demonstrates that LDA can be used to filter and identify messages with some consistent and homogenous meaning. However, several topics had more than one significant intent. This finding is not surprising given that patients and providers alike may need to provide context for information requests or summative updates using the program, e.g., initiating communication with a pleasantry, providing BP measurements, describing BP cuff or program error, and reporting medication taking patterns. Moreover, when we set out to validate the LDA-derived topics via manual annotation, we still found significant heterogeneity within LDA-derived topics.

As a first step toward deeper intent classification, we manually annotated high-value, pre-defined medication intents with a fixed number of topics for medication-related messages by patients and providers (4 defined topics each). Given our manually-annotated reference standard, we observed that the distribution of messages with a single medication intent are largely skewed within both the patient-generated and provider-generated medication messages. We aimed to determine how well medLDA could identify these high-value medication intents as distinct topics. We observed that most patient-generated medication intents are *medication request* or *medication taking* which tend to co-occur among medication LDA topics. Among the provider-generated medication intents, *medication question* was predominant (∼70%) among messages. Among the grouped patient medication messages, 3 of 4 medication intents were observed among medLDA topics. Among the grouped provider medication messages, 2 of 4 medication intents were observed among medLDA topics. We suspect that these skewed distributions and shared common vocabulary terms may explain why the medLDA models were not able to perfectly discern each medication intent across topics. Further investigation of terms and semantic categories encoded by MedEx and STANZA provided some insights of shared and distinct concepts; however, more powerful language models might be necessary to discern intents with subtle semantic differences that are important for clinical contexts.

### Limitations and Future Work

Our pilot study has several limitations. Notably, we conducted our analysis with a single dataset generated from a particular patient-provider engagement program. However, we believe that unsupervised learning approaches can be beneficial for streamlining the mining of free-text data with customization to each individual program or application. As a result, this work is important because it highlights potential approaches for incorporating unstructured learning into this process. Customizations could be achieved with a larger corpus and more powerful language models. In the future, we will develop patient-provider language models such as Bidirectional Encoder Representations from Transformers (BERT) models that might improve our ability to capture and leverage differences between message types to improve automatic intent classification for medications and other high-value intents.

## Conclusion

We demonstrated how unsupervised learning can be applied to group text messages and identify medication-related messages within a bidirectional text messaging system for hypertension management. While LDA was useful in generating coarse categories, more detailed intent annotation is needed to develop reliable NLP-based intent classifiers that drive clinical actions and address subtopic heterogeneity.

## Data Availability

Summary data files will be available at https://github.com/semantica-NLP/LDA_textbot_analysis

## Acknowledgement

The project was supported in part by grant number P30-AG034546 from the National Institute on Aging which provided financial support for Anahita Davoudi, Thaibinh Luong, Timothy Delaney, and Elizabeth L Asch. Krisda Chaiyachati reported receiving grant support from Independence Blue Cross, Inc. for this work. Danielle Mowery was funded by the National Institutes of Health UL1-TR001878 for this work.

## Author’s Contributions

AD, NL, TL, KHC, EA, and DM conceptualized the study design. TL and NL provided and helped interpret data. DM, NL, and TL coded text data, and AD analyzed the data. AD and DLM created the data visualization. DLM, AD wrote the first draft of the manuscript. All authors revised the manuscript and approved the final version.

